# Regional patterns and treatment-seeking behaviours for hypertension in Lesotho: Analysis of Lesotho Demographic and Health Surveys (DHS) 2023 data

**DOI:** 10.1101/2025.03.20.25324304

**Authors:** Vishal Tikhute, Balaka Chattaraj

**Affiliations:** Research and Development Wing, Pragati Creations, Pune, India; Social Work department, Tezpur University, Tezpur, India

**Keywords:** Cross-Sectional Studies, Prevalence, Lesotho, Hypertension, Healthy Lifestyle, Delivery of Health Care

## Abstract

Attributed to poverty and resource constraint situations, the population in low- and middle-income sub-Saharan countries like Lesotho is at high risk of preventable non-communicable diseases such as hypertension. Literature showed a high burden of hypertension in sub-Saharan Africa. However, no study assessed the burden of hypertension and the situation of treatment-seeking among the household population of Lesotho.

**Objectives:** The study aimed to assess the regional patterns of hypertension and treatment-seeking among the household population of Lesotho.

**Methods:** We analysed recently released Lesotho DHS 2023 data and presented district- and ecological zone-wise prevalence of hypertension and treatment-seeking among the Lesotho household population.

**Results:** The prevalence of hypertension among the household population was 1.5% (95% CI 1.40-1.66). Prevalence was highest in Maseru district (2.4%; 95% CI 1.90-2.90) and lowest in Thaba-Tseka district (0.6%; 95% CI 0.37-0.86). The treatment-seeking among diagnosed individuals in Lesotho was 66.8% (60.21-74). Thaba-Tseka district (50%) and capital city Maseru (51%) had the lowest number of patients who took treatment for hypertension.

**Conclusions and Recommendations:** The prevalence of hypertension is high among the Lesotho household population. Further, despite treatment being provided free of charge, every one out of three patients don’t take antihypertensive treatment. Despite high prevalence, the treatment-seeking is lowest in the capital city, Maseru. Priority attention should be given to districts such as Maseru, Mafeteng, Quthing, Qacha’s Nek, and Thaba-Tseka, where treatment-seeking is low. This highlights the need for revising the existing healthcare delivery mechanism and prioritising community needs in health policy. Community-based interventions promoting healthy lifestyles and periodic monitoring were recommended.

## 1. Introduction

There is a high burden of hypertension in the Sub-Saharan African region (including Lesotho) [1]. As per WHO, for Lesotho, the age-standardised prevalence of hypertension among adults aged 30–79 years for the year 2019 was 40%, which is higher than the global prevalence of 32% [2]. High salt intake, tobacco use, obesity, alcohol consumption, and physical inactivity contribute to a high hypertension burden in Lesotho (WHO, 2024). Hypertension in the country is closely associated with high morbidity and mortality due to its complications, such as cardiovascular diseases [2], [3]. Despite a high number of hypertension cases reported in Lesotho, many other cases are still underdiagnosed; people suffer from a lack of access to quality health care and unawareness about the problem [1].

Controlling hypertension requires a series of sequential steps. The steps to initiate treatment are screening of patients, creating awareness about treatment, treatment initiation, and long-term retention and treatment control [1], [4]. However, literature suggests that in Lesotho, health care services primarily focused on diseases related to infection and maternal health [5]. Geographical limitations and regional variations that might be detrimental to treatment-seeking for hypertension have been scarcely described in the literature [6]. However, no study has used a recently released nationally representative sample from DHS and presented the actual magnitude of hypertension and regional distribution of treatment-seeking for hypertension among the Lesotho household population. Therefore, it is crucial to study the regional patterns and treatment-seeking behaviours for hypertension in Lesotho by analysing the most recent Lesotho Demographic and Health Surveys (DHS) 2023 data.

### 2. Methods

The Lesotho Kingdom has ten administrative districts, that are classified into four ecological zones, namely the lowlands, foothills, and highlands (mountains) and the Senqu Valley [6], [7]. A cross-sectional study design was used to present the district and zone wise regional patterns of hypertension and treatment-seeking among the Lesotho household population. We analysed secondary data from the household member recode dataset (LSPR81FL.DTA file) of the Lesotho DHS 2023 [8]. This is the most recent round of DHS, which covered a total population of 36,161 household members from 10 districts of Lesotho. We used these entries to present district- and zone-wise prevalence of hypertension in Lesotho.. We used these entries to present district- and zone-wise prevalence of hypertension in Lesotho. Further, a total of 555 individuals who had been diagnosed (pre-diagnosed cases) by healthcare professionals as hypertension patients were selected as a sample to present the regional pattern of the treatment-seeking among ten districts and four ecological zones of Lesotho. To assess the prevalence, the dichotomous variable ‘ever been diagnosed by a doctor/health worker with hypertension’ (response yes or no) was considered for analysis. Similarly, for the treatment-seeking variable, ‘Has a doctor/health worker prescribed medication to control blood pressure?’ (response yes or no) was considered. Descriptive statistics were used to analyse DHS data and to present the district- and zone-wise prevalence of hypertension and treatment-seeking in Lesotho.

## 3. Results

The prevalence of hypertension among the Lesotho household population (N = 36,161) was 1.5 percent (95% CI = 1.40-1.66). Prevalence was highest in Maseru district (2.4%; 95% CI 1.9-2.9) and lowest in Thaba-Tseka district (0.6%; 95% CI 0.37-0.86). Four districts, Maseru district (2.4%), Berea (2.4%), Butha-Buthe (1.8%), and Mafeteng (1.7%), had prevalence higher than national prevalence (Table 1).

**Table 1.** Prevalence of hypertension in ten districts and four ecological zones of Lesotho (*N* = 36,161)

| Area | n | % | Total |
| --- | --- | --- | --- |
| **District** |  |  |  |
| Butha-Buthe | 64 | 1.8 | 3,612 |
| Leribe | 59 | 1.4 | 4,219 |
| Berea | 92 | 2.4 | 3,886 |
| Maseru | 96 | 2.4 | 4,014 |
| Mafeteng | 57 | 1.7 | 3,408 |
| Mohale'S Hoek | 45 | 1.4 | 3,133 |
| Quthing | 47 | 1.4 | 3,405 |
| Qacha'S Nek | 39 | 1.3 | 2,964 |
| Mokhotlong | 32 | 0.9 | 3,387 |
| Thaba-Tseka | 24 | 0.6 | 4,133 |
| **Ecological Zone** |  |  |  |
| Lowlands | 348 | 2.1 | 16,766 |
| Foothills | 35 | 1 | 3,412 |
| Mountains | 99 | 0.9 | 10,557 |
| Senqu River Valley | 73 | 1.3 | 5,426 |
| **Lesotho (Total)** | **555** | **1.5** | **36.161** |

Further, considering the ecological zones, the prevalence was highest in lowlands (2.1%), followed by the Senqu River Valley (1.3%), foothills (1%), and mountains (0.9%). The mean age of the sample population (*n* = 555) was 38.3 (SD = 9.9). One third were males. Eighty-four percent were ever-married individuals. The majority were educated (97%). Fifty-six percent lived in rural areas. Sixty-three percent lived in lowlands, followed by mountains (18%), the Senqu River Valley (13%), and foothills (6%). Twenty-seven percent belonged to the richest wealth index (Table 2).

**Table 2.**
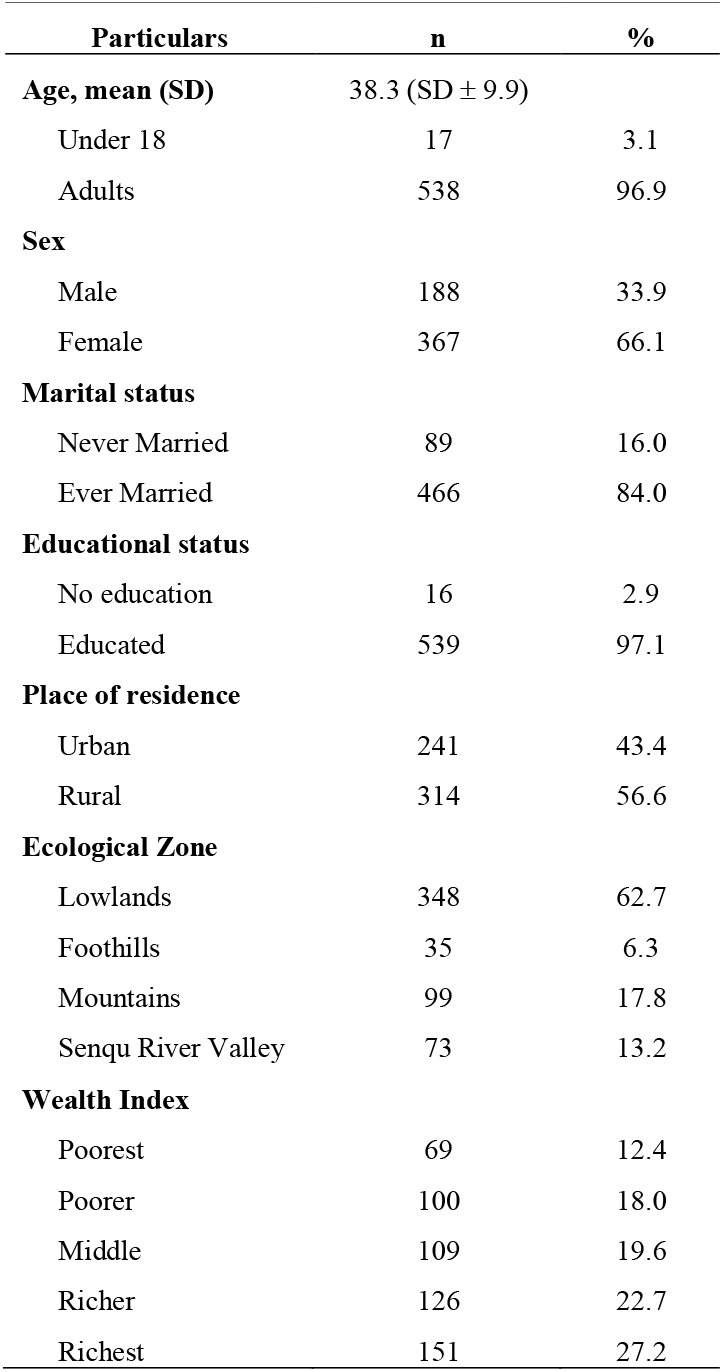
Socio-demographic characteristics of study sample (*n* = <u>555)</u>.

Sixty-seven percent of individuals reported that healthcare professionals had prescribed medicines to lower their blood pressure. The treatment-seeking among diagnosed individuals was 66.8% (60.21-74). Mokhotlong district (93.8%) had the highest proportion of patients that took treatment for hypertension, while it was lowest in Thaba-Tseka district (50%). Five districts, i.e., Maseru (51%), Mafeteng (64.9%), Quthing (55.3%), Qacha’s Nek (51.3%), and Thaba-Tseka, had lower proportions of patients who took treatment than the national prevalence. With the exception of Mafeteng, the rest of these four districts had populations living in mountainous regions (Table 3).

**Table 3.** Treatment seeking among hypertension patients in Lesotho (*n* = 555)

| Area | Prescribed<br>medicine<br>(%) | Taking<br>treatment<br>(%) | N |
| --- | --- | --- | --- |
| **District** |  |  |  |
| Butha-Buthe | 60.9 | 87.5 | 64 |
| Leribe | 76.3 | 89.8 | 59 |
| Berea | 58.7 | 62.0 | 92 |
| Maseru | 67.7 | 51.0 | 96 |
| Mafeteng | 71.9 | 64.9 | 57 |
| Mohale'S Hoek | 82.2 | 68.9 | 45 |
| Quthing | 66.0 | 55.3 | 47 |
| Qacha'S Nek | 59.0 | 51.3 | 39 |
| Mokhotlong | 78.1 | 93.8 | 32 |
| Thaba-Tseka | 58.3 | 50.0 | 24 |
| **Ecological Zone** |  |  |  |
| Lowlands | 67.8 | 66.7 | 348 |
| Foothills | 60 | 85.7 | 35 |
| Mountains | 63.6 | 58.6 | 99 |
| Senqu River Valley | 74 | 69.9 | 73 |
| **Total** | **67.4** | **66.8** | **555** |

## 4. Discussion

We have presented the regional distribution of hypertension and treatment-seeking among hypertensive patients in the sub-Saharan country of Lesotho. This is the most recent assessment that presented the nationwide situation of hypertension and treatment-seeking among hypertension patients. The earlier studies have highlighted the high prevalence of hypertension in Lesotho [1], [2], [9]. Some of these studies have linked age and behavioural factors with the high burden of hypertension among citizens of Lesotho [9], [10]. However, the situation of treatment-seeking for hypertension was not explored before. Further, an earlier study that used an old DHS dataset (from the year 2014) also presented similar findings; however, this study had assessed prevalence among women of childbearing age only (15–49 years) [10]. All these earlier studies had only covered specific age groups (15-49 years), rural populations, and women of childbearing age only [10]; and other age groups, urban residents, and males were excluded. Also, Lebuso and De Wet-Billings (2022) had used an old DHS dataset from the year 2014, while we used the most recent 2023 dataset. Further, the prevalence of hypertension among women mentioned in their study was higher than what we have calculated using DHS 2023 data, suggesting the possibility of improvement in the situation of hypertension among Lesotho households during the last ten years. Similar to our findings, this study had also reported higher prevalence among the married population. Further in alignment with this study’s findings, we found that the Maseru district, where the capital of Lesotho is situated, had the highest prevalence among all ten districts of Lesotho.

Our results also differ from the prevalence presented by the WHO for Lesotho (40%), as they presented prevalence for the year 2019, and their assessment only covered adults aged 30–79 years [2] while we have covered all age populations (15 years and above). In Lesotho, services for hypertension and diabetes are predominantly free in public health centres, and patients are only required to pay a minimal fee when attending hospital services [1]. Despite this, we found that one third of hypertension patients did not take treatment. The possible justification for this can be the several challenges that have been discussed elsewhere [6], [11], [12], [13]. That includes fragmented service delivery, geographical limitations, dependency on external donations, lack of accountability, and no priority to preventable health conditions, such as hypertension [6], [11], [12], [13]. The consequences of separate, specialised services undermine holistic, individualised patient care, patients’ adherence to medication, and multiple clinic visits, each time enduring long waiting periods, and ultimately discontinuation of treatment [6], [14]. Resultantly, despite the availability of highly specialised care, the burden of hypertension and other preventable health conditions is rising in Lesotho [6], [12], [13].

Further, Lesotho is classified as a lower-middle-income country, and 57.1% of the population lives below the national poverty line [6], [15]. Poverty is particularly acute in the mountainous areas, which are difficult to reach [11]. In alignment with this, we found that the treatment-seeking was low in mountainous regions of Lesotho. In Lesotho, the treatment of hypertension is guided by the national guidelines on the management of diabetes and hypertension [6]. Community health workers, also known as village health workers (VHWs), serve as the key foundation of healthcare delivery in Lesotho [6]. A recent cohort study demonstrated the efficacy of community-based hypertension management through the VHWs channel [16]. They found that community-based management of hypertension through VHWs effectively improved treatment-seeking, treatment compliance, and better health outcomes in later stages [16]. The model can be effective in mitigating geographical challenges in the provision of services for hypertension management. As in the current study, we found that the mountainous regions of Lesotho had low treatment-seeking for hypertension. This points out the challenges experienced by hypertension patients accessing treatment due to diverse geographical (mountainous topography) and harsh weather conditions [6]. Considering all these challenges, our study has provided important insights and evidence that can be further used to strengthen healthcare service delivery, particularly for preventable health conditions such as hypertension.

## 5. Conclusions and Recommendations

The prevalence of hypertension is high among the Lesotho household population. However, treatment-seeking (i.e., individuals taking antihypertensive drugs to lower their blood pressure) varies across districts and ecological zones of Lesotho. Despite the availability of speciality care, one out of every three hypertension patients don’t take antihypertensive drugs to lower the blood pressure. Urbanisation has significantly contributed to the high burden of non-communicable diseases such as hypertension in Lesotho. Urban areas have a high prevalence, almost twice that of rural areas. Further, the regional distribution of hypertension highlights the need to prioritise service delivery in highly urbanised areas, such as the capital city of Maseru, which has the highest burden of hypertension. Lowlands and the Senqu River Valley have a high burden of hypertension.

While for treatment seeking, mountainous ecological zones have the lowest proportion of individuals who take antihypertensive treatment. This suggests a detrimental effect of geographical barriers and harsh weather conditions on treatment-seeking for hypertension among the Lesotho household population. To address these issues, there is a need for community-based interventions that can supplement efforts by the existing healthcare services to reduce the burden of hypertension in the country. In Lesotho, particularly in rural settings, VHWs can offer lifestyle counselling, basic guideline-directed antihypertensive medications, lipid-lowering, and antiplatelet treatment supported by a tablet-based decision support application to eligible participants [16].

### Public Health Implications

- Despite the availability of speciality care, Lesotho has a high burden of hypertension.
- Geographical and regional distribution of hypertension highlights the need for a focused approach to control hypertension in Lesotho. The capital city of Maseru, lowlands, and mountainous regions have a high prevalence of hypertension. These areas need priority attention from policymakers, where special focus should be given to improving service delivery, ultimately supporting treatment-seeking for hypertension among the Lesotho household population.

## Data Availability

The data analysed in this study can be requested from The DHS program at https://dhsprogram.com/Data/terms-of-use.cfm

## Author Declarations

## Ethics approval and consent to participate

Data privacy and ethics followed as per the DHS data privacy statement available at https://dhsprogram.com/Data/terms-of-use.cfm. Ethics approval from the IRB does not apply to this study. The study is based on anonymous secondary data available only for the registered research studies at the DHS data portal (i.e., only after obtaining authorisation). The dataset available on the DHS portal is state-level collective data and does not include individual/personal information. Also, the study is not a part of any clinical trials and doesn’t involve human/living subjects/animals. Therefore, IRB approval is not needed for this study. The concerned agencies (i.e., the Ministry of Health, Lesotho; The DHS Program; and ICF) and sources have been cited in the reference section. Upon obtaining written permission, DHS allows the use of this data (downloaded from the DHS data portal) for registered research studies by researchers. The authors/researchers of this study have obtained the authorisation letter from the DHS program, allowing the use of Lesotho DHS 2023 data for this study. Obtaining individual consent is also not applicable, as the study used anonymous secondary data.

## Acknowledgements

We are grateful to the USAID-Demographic and Health Surveys (DHS) Program, Rockville, Maryland, USA for authorising the use of the Lesotho DHS 2023 dataset for this study.

## Author contributions

VT conceptualised and designed the entire study. VT also drafted the primary draft of the manuscript, revised it, and finalised it for submission. VT was associated with the responsibility of approaching data sources, obtaining authorisation from the DHS program, data cleaning, data analysis, and presenting findings. BC drafted the primary draft of the introduction section. BC also reviewed the final manuscript.

